# Trajectory-Informed Breathomics for Dynamic Mapping of Health and Disease: Toward a *Health Navigation Framework*

**DOI:** 10.1101/2025.09.01.25334816

**Authors:** Kenta T. Suzuki, Atsuhiro Nagasaki, Shinnichi Sakamoto, Kazuhisa Ouhara, Hideyuki Hyogo, Hiroshi Aikata, Toshinobu Takemoto, Akiko Tanaka, Goro Funakoshi, Yutaro Koyama, Kazushi Ikeda, Woosuck Shin, Kazuo Sato, Takashi Takata, Yuichi Sakumura, Mutsumi Miyauchi

## Abstract

**Background:** Most diagnostic frameworks treat disease as static, overlooking its inherently dynamic and continuous progression. Noninvasive biomarkers capable of capturing physiological trajectories are critically needed for proactive health management.

**Methods:** We developed a breath-based health navigation framework using exhaled volatile organic compounds (VOCs) as integrative, real-time indicators of systemic metabolism and immunity. Breath profiles were collected from healthy individuals and patients with nonalcoholic steatohepatitis (NASH, recently redefined as metabolic dysfunction–associated steatohepatitis, MASH), hepatocellular carcinoma (HCC), and periodontitis (PD). Multidimensional analysis and trajectory-informed mapping were applied to project individual health states into a low-dimensional physiological space.

**Results:** Ratio-normalized VOC profiles revealed disease-specific metabolic signatures across P450-derived and microbiota-derived compounds. Machine learning achieved high diagnostic accuracy for distinguishing health, NASH, HCC, and PD, while dimensionality reduction and topological analysis visualized a continuous progression from health to advanced liver disease. This approach captured preclinical shifts and transitional states often missed by static diagnostics.

**Conclusions:** Exhaled VOCs can serve as dynamic biomarkers for mapping health–disease transitions. By offering clinicians an intuitive map to locate patients within the health–disease continuum and anticipate their trajectories, this framework enables proactive decision-making and personalized intervention strategies. Collectively, our work points toward a paradigm shift in disease monitoring, with future integration into portable sensing technologies for noninvasive, continuous health-state assessment.

## INTRODUCTION

Modern diagnostic frameworks in medicine remain largely dependent on static, categorical classifications of disease, often overlooking the inherently dynamic and continuous nature of physiological deterioration ^1^. This structural limitation hinders timely intervention and prevention, especially in the context of chronic diseases ^2^, by obscuring early, subclinical transitions.

Metabolic dysfunction-associated steatotic liver disease (MASLD) exemplifies such challenges. MASLD arises in association with obesity, type 2 diabetes, and other metabolic disorders, progressing from simple steatosis to metabolic dysfunction–associated steatohepatitis (MASH; formerly termed nonalcoholic steatohepatitis, NASH), fibrosis, cirrhosis, and ultimately hepatocellular carcinoma (HCC) in a multi-stage manner ^3–5^. This disease trajectory is influenced not only by hepatic metabolic dysfunction but also by gut microbiota alterations ^6^, the gut-liver axis, and systemic inflammatory conditions such as periodontitis ^7^, operating through metabolic–immune– microbiome crosstalk. MASLD thus represents a networked, multi-organ pathology that resists adequate characterization by static snapshots of single-organ function ^8^.

Current diagnostic tools, including blood tests, imaging, and molecular markers, are often based on single time-point measurements and lack the temporal resolution to capture subtle physiological transitions ^9^. In contrast, volatile organic compounds (VOCs) in exhaled breath offer a portable, noninvasive, real-time window into systems-level metabolism. VOCs are generated through endogenous metabolic and detoxification processes involving the liver, gut microbiota, kidneys, immune system, and other organs, and are excreted via the lungs ^10,11^. Breath VOCs are particularly informative in liver diseases, where specific ingerprints of aldehydes, ketones, and short-chain fatty acids have been shown to correlate with disease state ^12,13^ and may anticipate clinical change. These compounds often capture complex, multisystem perturbations that are inaccessible via blood or urine biomarkers ^11,14^, including cross-organ interactions.

Recent studies have demonstrated the potential of breath VOCs in diagnosing liver diseases such as cirrhosis or MASLD, often using classification models to distinguish disease from health ^13–15^. Yet, such binary, snapshot-based approaches do not address the more fundamental question of how physiological states evolve over time. They offer no information about directionality, transitional states, or pre-symptomatic trajectories or early warning signals.

Here, we propose a time-resolved *health navigation model* that maps individual VOC profiles— collected from healthy controls, patients with MASH, HCC, and periodontitis—into a topologically coherent two-dimensional metabolic state space. Using time-series data, this model visualizes physiological transitions as directional velocity vectors. Inspired by RNA velocity approaches ^16,17^, we adapt the notion of state-specific kinetics to VOCs dynamics so that the framework captures not only the position of an individual within the health-disease continuum but also the trajectory and momentum of physiological change.

This dynamic mapping system is envisioned not as a mere classification tool; it is designed to serve clinicians as an intuitive visual interface that reveals in near real time whether a patient is approaching or moving away from a specific disease state. In principle, such a framework could allow early detection of presymptomatic micro-transitions in physiological state—often preceding clinical symptoms or lab abnormalities—and may ultimately support personalized decisions about when to intervene.

Looking ahead, integration with simplified, real-time breath VOCs sensing devices could enable longitudinal, at-home monitoring in everyday environments, delivering actionable trajectory feedback to individuals and establishing a foundation for proactive, individualized health management.

## RESULTS

### Ratio-based VOC profiles reveal disease-specific metabolic signatures

Interindividual variability in exhaled VOC concentrations arises from several factors, including age, sex, smoking habits, and pulmonary filtering capacity. As shown in **Table 1**, the participants in this study—exclusively Japanese—displayed heterogeneous backgrounds, including differences in age and smoking status. While weak correlations were observed between some VOCs (such as o-xylene and p-xylene) and age (**Table S4**), these compounds exhibited low concentrations across all nonhealthy groups and did not display age-dependent trends within the healthy group (**Figure S1A**, **S1B**). Moreover, a summative analysis of 16 VOCs indicated a general decline in VOC levels with age and disease (**Figure S2A**, **S2B**), but considerable variation remained even among healthy participants. These findings underscore the necessity of normalizing VOC data to minimize the influence of lung function and individual variability.

**Table 1.** Participant characteristics. N/A represents the number of participants who did not give their consent to provide information on age and sex; their data were thus treated as missing.

| Disease | Sample | Age (years) | Smoking | Sex |  | N/A |
| --- | --- | --- | --- | --- | --- | --- |
|  |  |  |  | Male | Female |  |
| Healthy | 101 | 24.5±6.73 | 7 | 34 | 55 | 12 |
| NASH | 67 | 60.0±10.8 | 10 | 41 | 25 | 1 |
| HCC | 95 | 70.7±10.0 | 15 | 52 | 27 | 16 |
| PD | 29 | 67.8±11.5 | 4 | 11 | 18 | 0 |

To account for these confounding effects, we applied ratio-based normalization to the 16 VOC concentrations in each sample (see **Methods**). To interpret these normalized VOC profiles in relation to underlying pathophysiology, we categorized the 16 VOCs into three biologically meaningful groups: cytochrome P450-metabolized compounds (P450), gut microbiota-derived compounds (Gut), and other compounds (Others), as summarized in **Table 2**. This categorization allowed us to examine disease-associated trends at the functional group level (**Figure 1A–C**).

**Figure 1.**
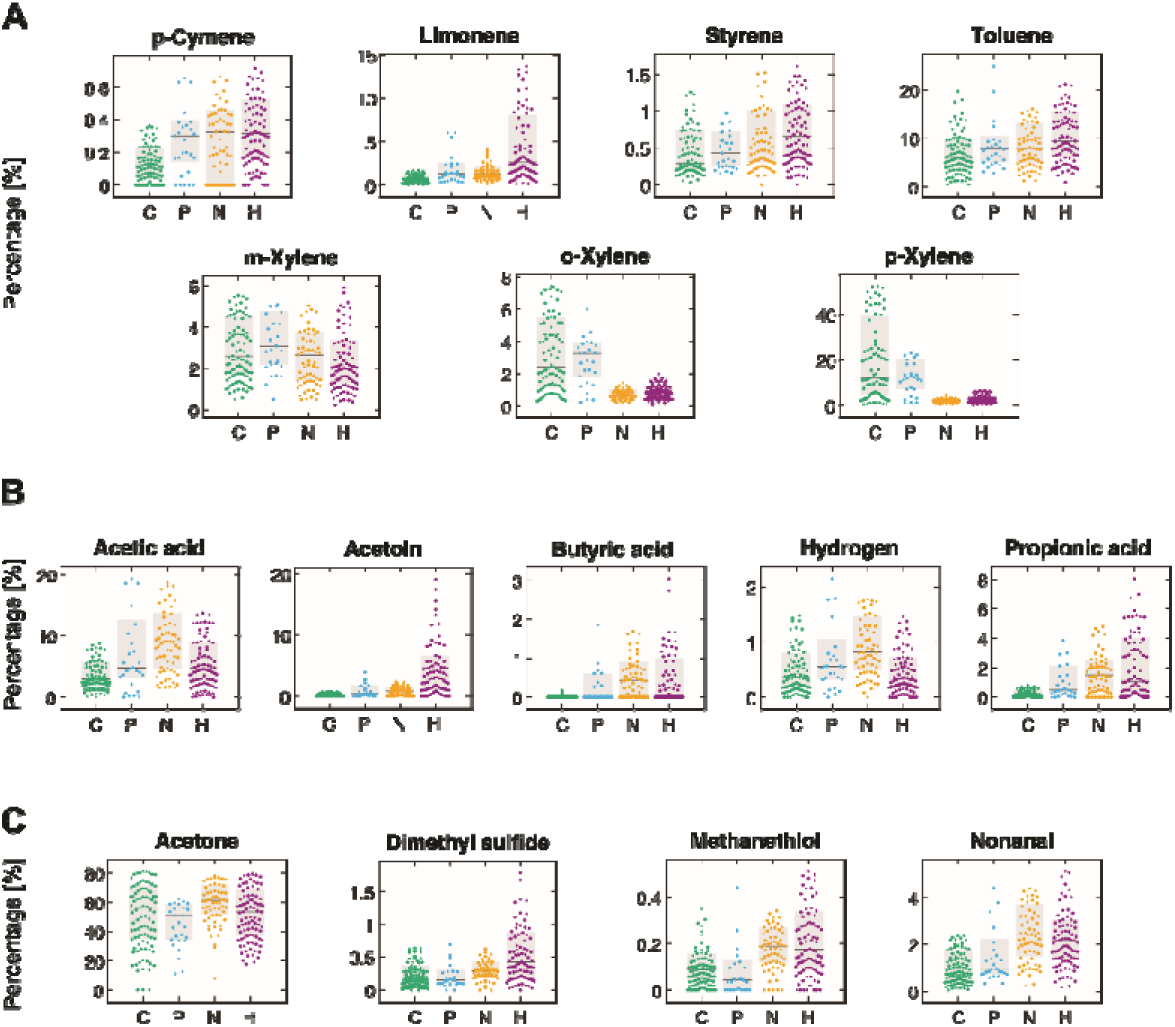
Distribution of VOC ratios in healthy and diseased patients highlighting differences in P450, gut-derived VOCs, and other VOCs. (A) Distribution of the percentages of VOCs metabolized by P450. (B) Distribution of the ratios of VOCs derived from gut bacteria. (C) Distribution of the proportions of other VOCs. C, P, N, and H represent the control, PD, NASH, and HCC groups, respectively. The boxplots show the median and interquartile range. For each participant, all 16 VOC ratios were obtained by dividing each VOC concentration by the sum of all VOC concentrations (total VOCs). Abbreviations: Ctrl, Healthy control; HCC, Hepatocellular carcinoma; NASH, Nonalcoholic steatohepatitis; PD, Periodontitis; VOCs, Volatile organic compounds.

**Table 2.** Categorization and breakdown of VOCs. P450-metabolized external components, bacterial gut components, and other components are abbreviated as P450. Gut, and Others, respectively.

| Category | Description | VOC |
| --- | --- | --- |
| P450 | Metabolized by cytochrome P450 in the liver and related to liver function. | p-Cymene, Limonene, Styrene, Toluene, (m, o, p)-Xylene |
| Gut | Mainly produced by gut bacteria or reflecting gut activity. | Acetoin, Acetic acid, Butyric acid, Hydrogen<sup>*</sup>, Propionic acid |
| Others | Identified in related studies. | Acetone, Dimethyl sulfide, Methanethiol, Nonanal |

<sup>\*</sup> Although hydrogen is not a VOC, it is included here as a metabolic product of gut bacteria.

In healthy individuals, the relative abundance of P450 and gut-derived VOCs was generally low, whereas these categories exhibited marked elevation in NASH and HCC samples. Among the 16 VOCs, no single compound served as a robust biomarker; however, disease-specific signatures appeared at the category level. Strikingly, xylene showed an opposite trend, with lower ratios in disease groups, reinforcing the importance of combinatorial assessment rather than reliance on individual VOCs. Moreover, acetic acid and acetoin showed opposite trends between NASH and HCC, while toluene and p-xylene also contributed to disease-specific patterns, especially in periodontitis (**Figure 1**).

To evaluate whether these category-based VOC ratios could differentiate health status, we constructed diagnostic models using supervised machine learning (see **Methods**). We tested three combinations of VOC categories: P450 alone, P450 plus gut-derived VOCs, and all three categories combined (**Figure 3**). As shown in **Figure 2** and **Table 2**, P450 VOCs alone achieved high classification accuracy, which improved upon inclusion of the gut-derived category (P450 + Gut). The addition of the third category (Others) offered no significant performance gain, suggesting that liver and gut-derived VOCs are the most informative. These results align with known pathophysiological changes in liver disease, such as cytochrome P450 dysfunction and altered gut flora ^6^. The stepwise improvement in classification performance when including Gut VOCs along with P450 VOCs was further validated in **Figure 2**, which illustrates the incremental contribution of each category to diagnostic accuracy.

**Figure 2.**
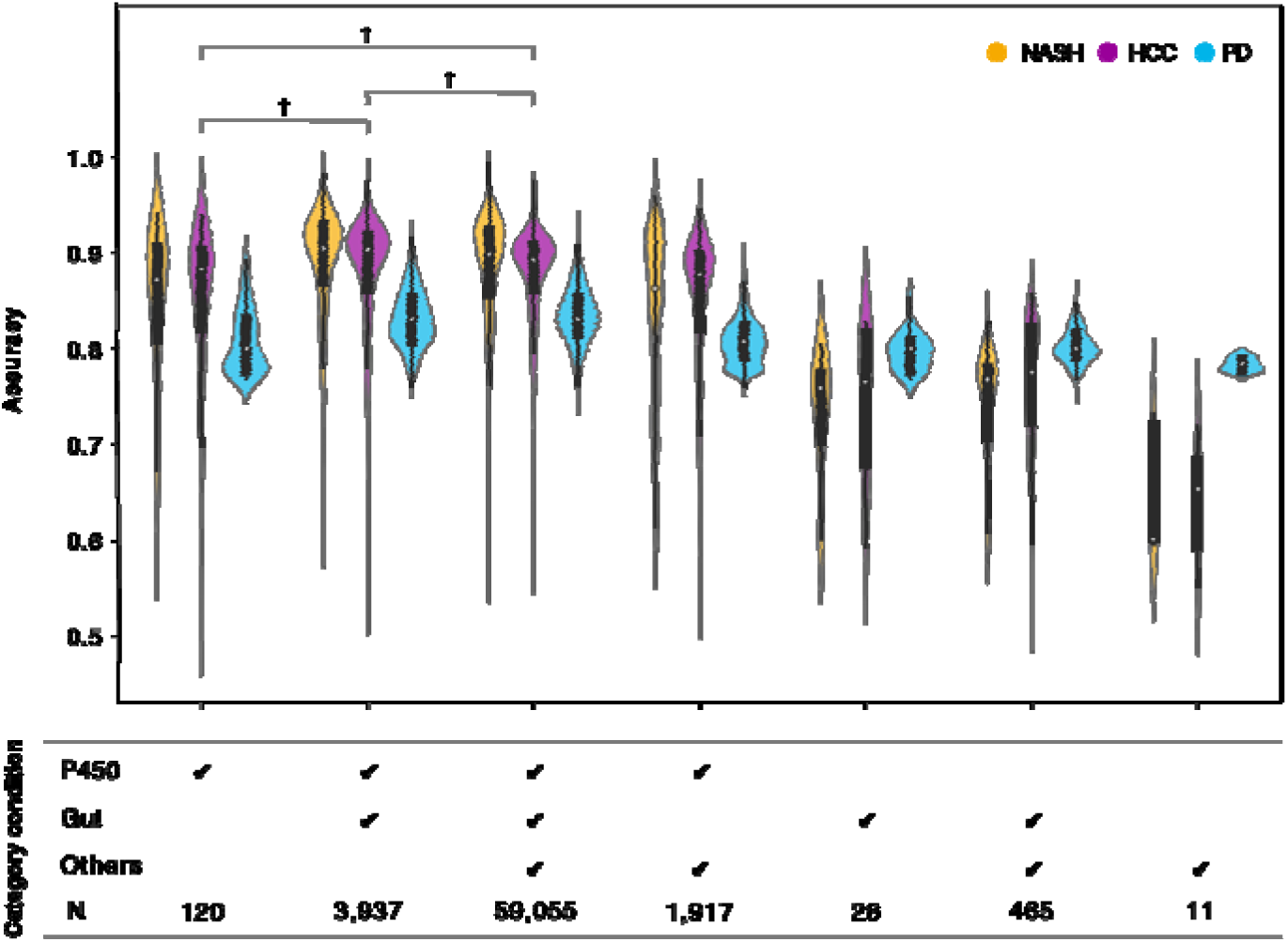
Accuracy of healthy vs. nonhealthy (i.e., disease) diagnoses for each combination of VOC categories. The horizontal axis represents the different combinations of VOC categories. Diagnostic accuracy was calculated for each combination using the corresponding set of VOCs. We employed ratios specific to selected VOCs (see Methods). For a single VOC category, the diagnostic accuracy is shown using only the VOCs within that category. Significance difference tests (†, Kruskal□Wallis test, significance level p = 0.01) were performed to compare the means among the three conditions on the left-hand side, and the results were significant for all three diagnoses. The mean values for diagnostic accuracy are provided in Error! Reference source not found., while **Suppl. Table 5** shows the maximum values for each precision group. Abbreviations: Gut, bacterial gut components; Others, components not included in P450 or the gut; P450, cytochrome P450-metabolized external components.

**Figure 3.**
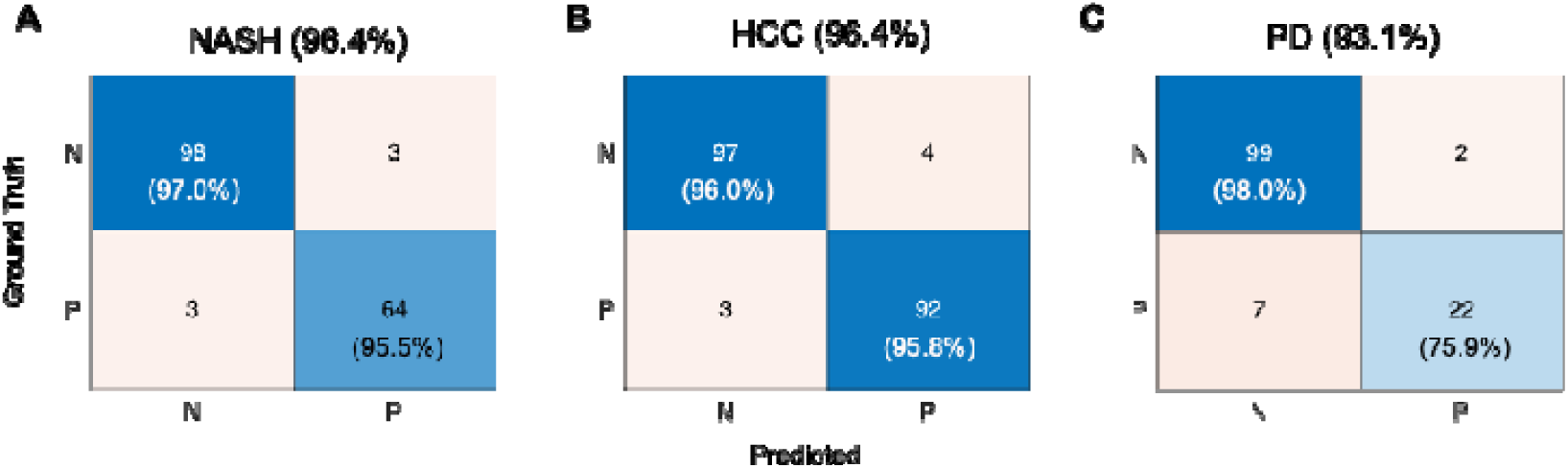
Diagnostic performance when using combinations of VOCs with the highest accuracy. Each confusion matrix demonstrates the diagnostic performance for (**A**) NASH, (**B**) HCC, and (**C**) PD patients relative to healthy controls. The rows denote the actual class, while the columns represent the predicted class. Diagonal elements represent correctly classified instances, whereas off-diagonal elements signify misclassified instances. The values in the title, top left, and bottom right represent accuracy, specificity, and sensitivity, respectively. Abbreviations: Ctrl, Healthy control; HCC, Hepatocellular carcinoma; N, healthy; NASH, Nonalcoholic steatohepatitis; P, disease; PD, Periodontitis.

Importantly, omitting the ratio normalization step led to a marked decline in diagnostic performance (**Figure S3**), confirming its critical role in extracting meaningful disease-related information. The detailed composition and distribution of these optimal VOC combinations are shown in **Figure S4**.

Together, these findings demonstrate that the combined use of VOC ratio normalization and functional categorization reveals disease-specific metabolic alterations. Rather than focusing on absolute concentrations or individual compounds, our results highlight the diagnostic value of category-level VOC patterns and establish a foundation for dynamic mapping of health–disease trajectories in subsequent analyses.

### Mapping health–disease transitions using exhaled VOCs

To move beyond binary classifications of “healthy” and “unhealthy,” we examined whether VOC profiles could support disease-resolved differentiation. While the high diagnostic accuracy achieved for each group (**Figure 2, Table 3**) suggests that VOCs encode meaningful health information, it remained unclear whether these patterns were disease-specific or simply reflected a general shift away from health. To address this, we analyzed the distribution of VOC ratios across multiple disease classes.

**Table 3.** Means and standard errors of accuracy by category condition in Figure 2. The bolded values indicate the highest accuracy for each disease.

| Category condition | NASH | HCC | PD |
| --- | --- | --- | --- |
| P450 | 0.85 ± 0.08 | 0.85 ± 0.08 | 0.81 ± 0.03 |
| P450, Gut | **0.89 ± 0.06** | **0.88 ± 0.06** | 0.83 ± 0.03 |
| P450, Gut, Others | 0.88 ± 0.06 | 0.88 ± 0.05 | **0.84 ± 0.03** |
| P450, Others | 0.84 ± 0.09 | 0.85 ± 0.07 | 0.81 ± 0.02 |
| Gut | 0.73 ± 0.06 | 0.74 ± 0.08 | 0.80 ± 0.02 |
| Gut, Others | 0.74 ± 0.06 | 0.77 ± 0.06 | 0.80 ± 0.02 |
| Others | 0.65 ± 0.06 | 0.64 ± 0.06 | 0.78 ± 0.01 |

We first confirmed that several VOCs displayed disease-specific signatures (**Figure 1**, **Table S6, Figure S5**). For example, acetic acid and acetoin exhibited inverse trends in NASH and HCC, while p-xylene and toluene contributed distinctly to PD versus NASH classification. These patterns were not apparent in raw concentration data but emerged clearly after ratio normalization. Such compositional contrasts provided the molecular basis for moving from binary to multi-class discrimination.

To further validate the disease specificity of VOC profiles, we also performed binary classification between each disease and the healthy control group. As shown in **Figure 3**, optimal VOC combinations using support vector machine (SVM) classifiers enabled highly accurate discrimination of NASH (96.4%), HCC (96.4%), and PD (93.1%) from healthy individuals. These results confirm that disease-associated shifts in VOC profiles are not only detectable but also diagnostically meaningful, particularly for liver-related conditions.

Using random forest models and exhaustive feature selection (wrapper method), we identified an optimal combination of 14 VOC ratios that achieved an overall classification accuracy of 80.1% (234/292) across four classes: healthy, NASH, HCC, and PD (**Figure 4**). Given the small sample size for PD, misclassifications tended to assign PD patients to either healthy or liver disease groups. However, when restricted to the three major classes (healthy, NASH, and HCC), sensitivities consistently exceeded 80%.

**Figure 4.**
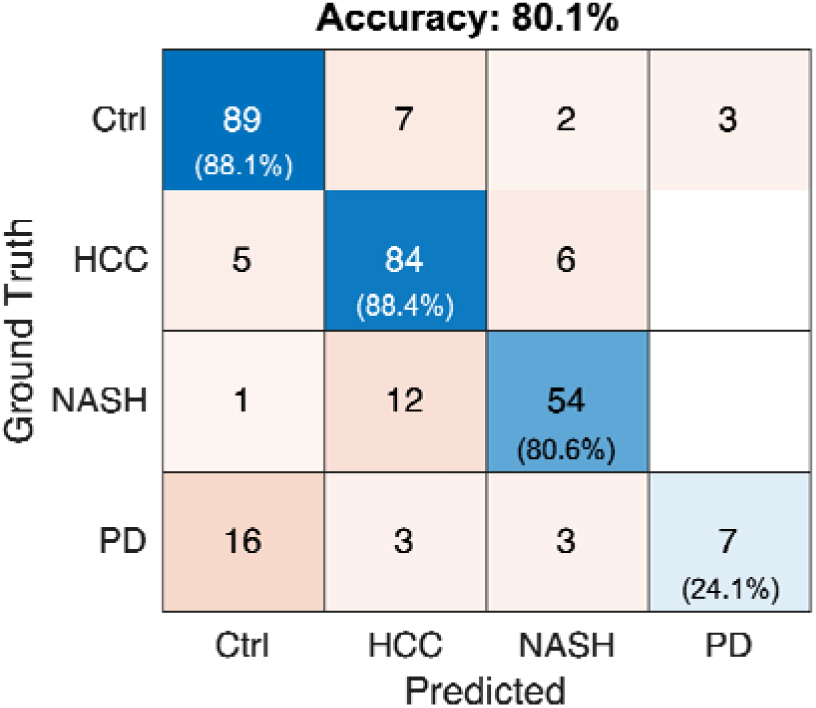
Four-class classification of healthy controls and patients with three diseases. The vertical axis shows the ground truth, indicating the true conditions, and the horizontal axis shows the conditions predicted by random forest classification.

Beyond classification, our primary objective was to visualize physiological transitions across disease states in a data-driven manner. To this end, we applied Uniform Manifold Approximation and Projection (UMAP) for dimensionality reduction and Partition-Based Graph Abstraction (PAGA) for topological trajectory analysis (**Figure 5**). The resulting two-dimensional map revealed distinct clusters corresponding to different health conditions. Healthy and PD samples were located at one end of the map, whereas HCC samples occupied the opposite side. NASH cases were broadly distributed between these two poles, suggesting a continuous trajectory from health to severe liver disease.

**Figure 5.**
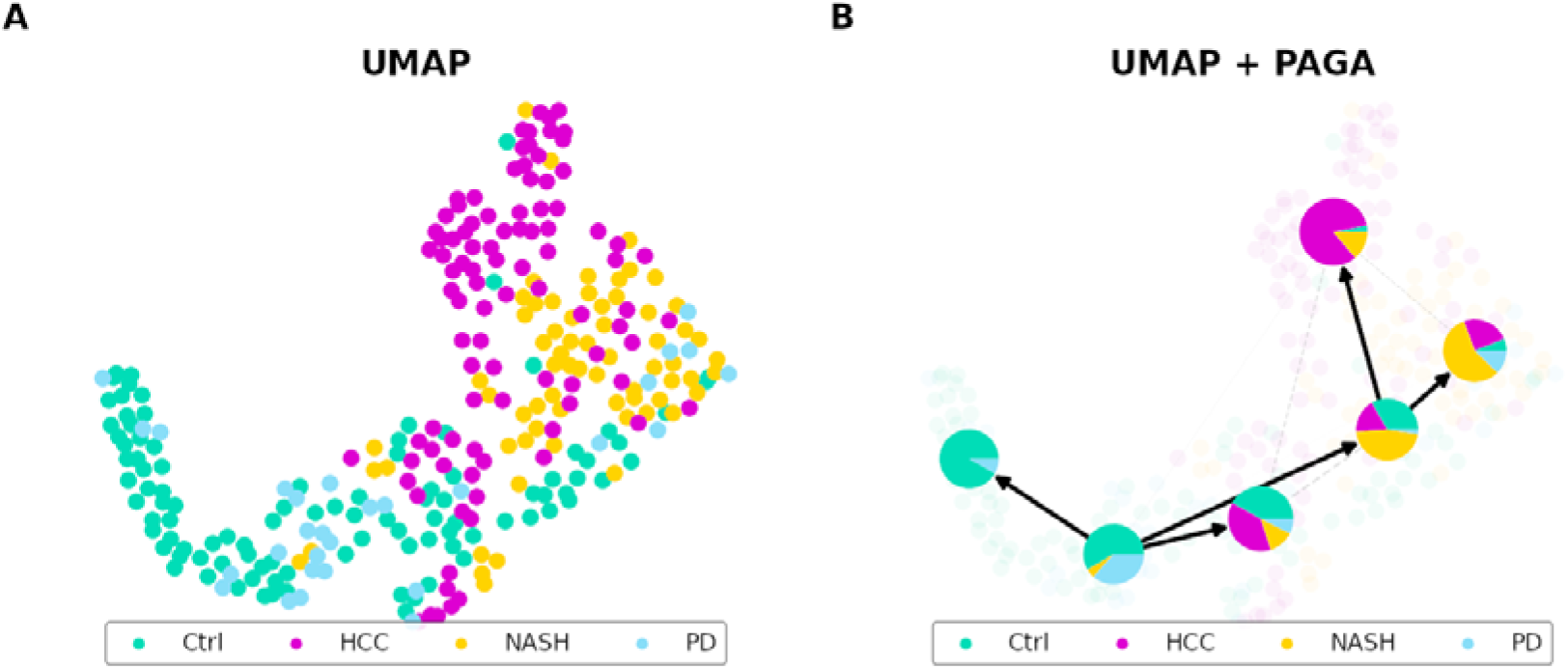
Visualization of disease transitions. (**A**) UMAP; for the 14 combinations of VOCs for which the highest accuracy was obtained in the random forest classification, the high-dimensional data were compressed to two dimensions using UMAP, a dimensionality reduction method. The data points belonging to the same cluster are plotted in close proximity. In the resulting embedding space, clusters were visualized using PAGA. The relationships between clusters are visualized as nodes and edges. Edges with weights greater than 0.55 remained. Green dots indicate healthy participants, blue dots indicate periodontitis patients, orange dots indicate NASH patients, purple dots indicate HCC patients, and white circles indicate nodes of PAGA-treated patients. (**B**) UMAP embedding with PAGA overlay. The same UMAP embedding is shown with a partition-based graph abstraction (PAGA) graph overlay, where PAGA represents the transition relationships between clusters depicted as nodes and edges. The pie charts overlaid on the nodes show the percentages of different classes within each cluster. Edges between nodes indicate the strength of connections between clusters, with thicker edges indicating stronger connections. The direction of the edges indicates the estimated direction of cellular transitions. Abbreviations: Ctrl, Healthy control; HCC, Hepatocellular carcinoma; NASH, Nonalcoholic steatohepatitis; PAGA, Partition-Based Graph Abstraction; PD, Periodontitis, UMAP, Uniform Manifold Approximation and Projection.

Importantly, these clusters preserved the topology of health states inferred from VOC composition, implying that spatial proximity and vector directionality in the two-dimensional map reflect underlying physiological transitions. This trajectory-informed map establishes a proof-of-concept *health navigation framework*—a visual interface for locating an individual within the health–disease continuum and anticipating the direction of physiological change.

## DISCUSSION

### Mapping Dynamic Health–Disease Transitions with Exhaled VOCs

Physiological states—including those of ostensibly healthy individuals—are not fixed but continuously drift along a multidimensional continuum ^1,2^. The direction and magnitude of these changes are often state-dependent and influenced by multiple overlapping biological processes. Capturing such transitions requires biomarkers that are not only sensitive and integrative but also amenable to frequent, noninvasive measurement. If such biomarkers could be mapped to reflect both current position and future trajectory within the health–disease landscape, they would provide a transformative tool for screening, monitoring, and clinical decision-making.

In this study, we demonstrate that exhaled VOCs—traditionally used in isolated disease contexts ^15,18^ —can serve a broader purpose when their compositional VOC profiles are analyzed in aggregate and normalized for interindividual variability. We propose a novel strategy to reduce confounding due to pulmonary function by preprocessing VOC concentrations into relative ratios. This normalization unlocked hidden disease-related structure, revealing category-level signatures that supported not only binary classification but also multi-class differentiation and trajectory visualization ^19^.

To construct a low-dimensional representation of this physiological landscape, we applied UMAP and PAGA—techniques originally developed to infer transcriptional trajectories in single-cell genomics^16,17^. When applied to our VOC data, these methods revealed a spatially ordered progression from healthy controls to NASH and finally to HCC (**Figure 5A**). The organization of PAGA nodes and edges further suggested substructures, possibly corresponding to distinct HCC etiologies, including metabolic (e.g., NASH) and viral origins (e.g., hepatitis B virus (HBV) or hepatitis C virus (HCV)) (**Figure 5B**). Thus, the topology of exhaled VOCs encodes not only diagnostic categories but also latent pathways of disease progression.

The high diagnostic performance achieved in this study—compared to prior reports ^20^ —was attributable to two key factors: the high detection rate of our VOC panel (**Table S1**), and our data preprocessing strategy based on ratio transformation. Certain compounds, such as butyric acid, were excluded from classification due to low detection frequency and information content, a critical step to avoid noise in predictive models ^21^. Without this ratio-based normalization, classification accuracy declined markedly (**Figure S3**), underscoring the importance of preprocessing. Although deep learning offers powerful alternatives for automated feature extraction, its prohibitive sample size requirements in clinical studies highlight the practical value of ratio-based approaches ^11^.

Although we focused on NASH and HCC in this analysis, the same approach could be extended to visualize transitions involving other diseases that share metabolic or inflammatory pathways. The VOC-derived coordinate space, once projected into a low-dimensional map, enables intuitive interpretation of patient trajectories across diverse disease states. Crucially, this method requires only breath samples and is compatible with real-time, continuous monitoring. This opens opportunities for applications where repeated blood sampling is impractical—such as intensive care, neonatal monitoring, and long-term at-home management.

Finally, unlike many previous studies that used VOCs primarily for classification without addressing underlying biological mechanisms ^18^, our approach provides a mechanistic rationale. The diagnostically informative VOCs identified here are metabolically linked to cytochrome P450 activity and microbial fermentation in the gut—two major systems implicated in chronic disease progression ^6,22^. In this sense, exhaled VOCs emerge not only as convenient diagnostic readouts but as dynamic windows into the integrative metabolic state of the body.

### Mechanistic Insights into Diagnostic Breath Biomarkers

To elucidate the biological underpinnings of the diagnostically informative VOCs identified in this study, we categorized them into three classes based on metabolic origin: P450-metabolized exogenous compounds, gut bacteria-derived metabolites, and other compounds (**Table 3**). Our results showed that combining VOCs across these categories substantially improved diagnostic accuracy for liver-related diseases (**Figure 2**). This integrative perspective highlights how multiple organ systems jointly shape the breath metabolome. Below, we examine the possible physiological mechanisms that may underlie VOC variations across different disease states.

### P450-metabolized VOCs

Several P450-metabolized compounds—including toluene, limonene, styrene, and xylene—were found to be useful in differentiating health states (**Table S6**, **Figure S5**). In particular, xylene concentrations were markedly lower in NASH and HCC samples compared to healthy controls and PD patients (**Figure 1A**). This depletion is consistent with impaired hepatic metabolism and clearance capacity, which redistributes the relative balance of exhaled VOCs. Xylene is metabolized in the liver into methylhippuric acid, a process modulated by toluene and other substrates competing for P450 enzymes ^23,24^. Disruption of this metabolic balance may thus explain the altered xylene levels observed. Furthermore, elevated toluene levels have been reported in patients with various cancers, including lung cancer ^25^, suggesting that dysregulated P450 metabolism may be a shared signature of malignancy. Although causal pathways remain unresolved, the consistent involvement of P450-related VOCs supports their potential as indirect markers of hepatic enzymatic activity. Given that cytochrome P450 expression is implicated in both liver disease progression and oncogenesis ^26^, these VOCs may represent accessible proxies of metabolic vulnerability.

### Gut microbiota-derived VOCs

We also observed disease-dependent differences of VOCs originating from gut microbial fermentation. Acetic acid and acetoin, in particular, varied across disease groups (**Figure 1**) and contributed strongly to HCC versus NASH classification (**Table S6**, **Figure S5**). Increased acetic acid in HCC patients may result from impaired hepatic clearance ^27^, whereas NASH patients may show elevated levels due to subclinical microbial ethanol production and subsequent metabolism ^28,29^. Acetoin, a microbial metabolite enriched in tumor microenvironments but scarce in healthy tissues ^30^, was especially useful in distinguishing HCC from NASH and PD. This tumor-associated specificity suggests that acetoin in exhaled breath could serve as a noninvasive prognostic marker for malignant transformation in NASH patients ^31^. While other bacterial VOCs such as butyric and propionic acid were not retained in the final model (**Table S5**), they may still hold relevance in broader disease contexts not examined here.

### Periodontitis-related mechanisms

Although PD is not a hepatic disorder, its inclusion provided insights into systemic disease interactions. VOCs associated with both P450 metabolism and gut microbiota contributed to PD classification (**Figure 2**). This aligns with prior evidence that chronic periodontal inflammation perturbs gut–liver axis and contributes to systemic diseases including diabetes and fatty liver ^32^. Periodontitis can disseminate bacteria, lipopolysaccharides, and cytokines into circulation, triggering hepatic inflammation and fibrosis ^33^. However, VOCs detected in deep lung exhalation are unlikely to originate directly from oral pathogens; rather, they represent systemically filtered metabolic products, distinct from volatile sulfides measured in oral headspace ^34^. Despite this limitation, we found that deep-lung VOC profiles enabled high diagnostic accuracy not only for PD but also for comorbidity detection (**Figure 2**, **Table 3**), underscoring their potential as holistic biomarkers of multi-organ crosstalk.

### Limitations and Future Directions

While this study demonstrates the diagnostic potential of exhaled VOCs and their utility in mapping physiological transitions, several limitations must be considered when interpreting the findings.

First, although we selected VOC combinations with the highest classification accuracy for each disease (**Tables S5** and **S6**), some combinations yielded closely comparable performance, making it difficult to determine whether the top-ranked sets were statistically superior. Additionally, certain compounds—such as p-cymene, limonene, acetoin, hydrogen, dimethyl sulfide, methanethiol, and nonanal—were associated with reduced classification accuracy for specific conditions like NASH (**Figure 2**), despite their presence in some high-performing combinations. These discrepancies suggest complex and possibly nonlinear relationships between individual VOCs and disease states. However, the primary goal of this study was not to evaluate negative predictors, but to identify configurations yielding high diagnostic accuracy. A more systematic evaluation of poorly performing combinations would require larger sample sizes.

Second, although our sample size exceeded those of previous VOC-based diagnostic studies ^20^, it remains insufficient for robust generalization, particularly for multigroup classification tasks involving smaller cohorts such as PD. Moreover, the underlying etiology of HCC (e.g., viral hepatitis vs. metabolic origin) could not be fully resolved owing to limited access to patient-level metadata. This limits our ability to validate whether distinct clusters observed in the UMAP and PAGA visualizations (**Figure 5**) reflect true etiological subtypes.

Third, diagnostic labels used in the machine learning pipeline were based on clinical diagnoses, which are inherently uncertain. While large datasets may help average out misclassification noise, future studies should consider incorporating longitudinal outcomes or biopsy-confirmed diagnoses to strengthen ground-truth labeling.

Fourth, standardization and reproducibility remain technical challenges. VOC concentrations may vary depending on instrument calibration, sensor type, and environmental factors. While we used ratio transformation to correct for interindividual variability in lung function, additional confounders—such as circadian rhythms, diet, medications, or age-related metabolic shifts—were not fully controlled. Similarly, our findings are derived exclusively from Japanese participants, whose gut microbiota and environmental exposures may differ significantly from other populations. Broader validation across racially and geographically diverse cohorts is therefore essential.

Despite these limitations, this study lays the foundation for a novel diagnostic framework based on breath analysis. The ability to project individuals—healthy and diseased—into a shared physiological space using a noninvasive, real-time biomarker opens avenues for proactive and longitudinal health monitoring. Once validated in larger and more diverse populations, this method could ultimately enable not only early diagnosis but also personalized intervention planning.

While this study employed GC–MS for high-resolution VOC detection, real-world implementation will require the development of portable, low-cost, and real-time sensing devices ^11,35^. Advances in compact semiconductor-based sensor arrays could enable widespread and on-demand VOC measurement, eliminating the logistical barriers of centralized laboratory analysis. Integration of such sensors into bedside monitors or wearable devices may transform this approach into a scalable and accessible *health navigation framework*, deployable in both clinical and everyday environments.

Collectively, this work points toward a paradigm shift: from discrete, episodic diagnoses to continuous, trajectory-informed navigation of health. Because everyone breathes, breathomics has the potential to democratize access to ultra-early, preventive diagnostics—allowing deviation from baseline health to be detected before symptoms arise, and ultimately contributing to reduced disease burden and healthcare costs.

## MATERIALS and METHODS

### Data Source

#### Study Participants

Exhaled breath samples were collected following approval by the Ethics Committee of Hiroshima University (approval number E-922). Written informed consent was obtained from all participants prior to sample collection. The study enrolled 67 patients diagnosed with nonalcoholic steatohepatitis (NASH), 95 patients with hepatocellular carcinoma (HCC), 29 patients with periodontitis (PD), and 101 healthy control participants (Ctrl). Participants were selected randomly and included only if they consented to participate. Given that diagnosis and enrollment occurred prior to the recent adoption of the MASH terminology, we use NASH throughout.

Breath samples were anonymized by removing personally identifiable information and assigning anonymized codes. Only non-identifiable demographic information, such as age and sex, was retained for analysis. Disease severity and stage were not recorded; each sample was labeled with a categorical disease group (NASH, HCC, PD, or Ctrl).

#### Definition of Disease Groups

At the time of data collection, NASH was diagnosed at the Department of Gastroenterology and Metabolism, Hiroshima University Hospital, according to the Japanese Society of Hepatology Guidelines ^36^. Diagnostic criteria included: (i) hepatic steatosis on imaging, (ii) alcohol intake <30 g/day (men) or <20 g/day (women), (iii) exclusion of other hepatic pathologies, and histological confirmation of steatohepatitis.

HCC was diagnosed according to the Japanese Society of Hepatology Guidelines for Hepatocellular Carcinoma ^37^, based on either histological confirmation or characteristic imaging findings. Underlying liver diseases in HCC patients included viral hepatitis (HBV or HCV), alcoholic liver disease, and NASH.

PD was diagnosed based on criteria from the Japanese Society of Periodontology ^38^, defined by the presence of periodontal pockets >4 mm with bleeding on probing, excluding cases of gingivitis.

The control group consisted of:

1. Healthy university students with periodontal pockets ≤3 mm,
2. Individuals with a history of periodontitis who had been successfully treated (current pockets ≤3 mm),
3. Individuals with pockets ≥4 mm but no bleeding,
4. University residents and staff with normal liver function tests.

All collected samples were included in the final analysis without exclusion.

#### Breath Sampling and VOC Measurement

Exhaled breath and ambient air samples were collected at Hiroshima University Hospital (Hiroshima, Japan) using 1 L polyvinylidene fluoride (PVDF) analytical barrier bags (Omi Odor-Air Service Corp., Omihachiman, Japan). To account for environmental background VOCs, ambient air within the sampling room was collected in parallel.

Prior to sample collection, participants were instructed to refrain from eating and smoking for at least two hours. They were seated in the breath sampling room for at least 10 minutes to equilibrate and breathe normally. Each participant then took two deep breaths, followed by a third deep breath with a 20-second breath hold. Exhalation was performed gently over ∼5 seconds into the barrier bag. Breath samples were collected from all participants after they received standardized training on the collection protocol. Personnel underwent supervised instruction and practiced until proficiency was confirmed, ensuring methodological consistency.

VOC analyses were performed within 32 hours of collection at the Knowledge Hub Aichi (Aichi, Japan). Two trained staff members conducted all collections for participants across all disease groups (Ctrl, NASH, HCC, PD).

#### VOC Analysis via GC–MS

VOC detection was conducted using a GCMS-QP2010 Ultra system (Shimadzu Corporation, Kyoto, Japan) equipped with a PY-3030D multifunctional pyrolysis unit (FRONTIER LAB, Fukushima, Japan). A capillary column (Ultra ALLOY-1, 60 m × 0.25 mm ID × 1.0 μm df; FRONTIER LAB) was employed for chromatographic separation.

Exhaled air from the PVDF bag was adsorbed onto TENAX TA or TENAX GR (GL Sciences Inc., Tokyo, Japan), then desorbed thermally using the PY-3030D system. Samples were cold-trapped using liquid nitrogen and analyzed with helium (purity 99.99995%; Taiyo Nippon Sanso, Japan) as the carrier gas. To eliminate ambient VOC contamination, background subtraction was performed based on previous methodology ^39^.

Hydrogen gas was analyzed separately using a thermoelectric hydrogen sensor-based breath analyzer ^40,41^, which incorporates a self-calibration feature and achieves ±2 ppm accuracy against a 200 ppm hydrogen standard gas.

A total of 16 compounds—including hydrogen (non-VOC)—were selected for downstream analysis based on high detection frequency across participants (**Table S1**). These compounds were categorized into three groups: (1) P450-metabolized external compounds, (2) gut bacteria–derived VOCs, and (3) other compounds, as defined in **Table 1**. The categorization was based on prior biochemical and epidemiological literature ^15,42^. Disease-specific associations of each compound are provided in **Table S2**.

### Analysis

#### Data Preprocessing

To address individual variability in exhaled VOC concentrations, we implemented a series of preprocessing steps. As a surrogate for pulmonary filtering capacity, we calculated the linear sum of all detected VOC concentrations. This approach leverages the law of large numbers to reduce individual variability, providing a robust estimate of lung function.

However, absolute concentrations of VOCs can be influenced by non-disease-related factors, potentially masking disease-specific signals. To mitigate this, we used relative concentration ratios rather than raw concentrations. Specifically, we computed the ratio of each VOC to the total concentration of VOCs that were deemed disease-relevant based on their diagnostic performance in machine learning models. This approach enhances sensitivity to disease-specific changes while minimizing noise from unrelated compounds.

#### Machine Learning–Based Classification

We employed supervised machine learning to classify participants by disease status using VOC concentration ratios. For binary classification tasks (e.g., healthy vs. diseased or between disease types), we used a nonlinear support vector machine (SVM) with a Gaussian kernel ^43^. The kernel width parameter (σ) and regularization parameter (C) were optimized via grid search to maximize classification accuracy (**Table S3**).

For multiclass classification (healthy, NASH, HCC, and PD), we used random forest models ^44^, which are well-suited for handling small-to medium-scale biomedical datasets. All analyses were conducted in MATLAB (R2023a; Statistics and Machine Learning Toolbox, MathWorks).

To identify diagnostically informative VOC subsets, we employed a wrapper-based feature selection method ^45^. We calculated classification accuracy for all possible combinations of two or more VOCs (excluding single-VOC models), resulting in a total of 65,319 evaluated subsets. These subsets were also categorized by their VOC type as defined in **Table 2** (P450-related, gut bacteria–related, or other), allowing for comparisons across biological pathways.

#### Accuracy Evaluation

Model performance was evaluated using 10-fold cross-validation ^46^. For each iteration, the dataset was split into 90% training and 10% testing sets. The testing outcomes were categorized as true positives, true negatives, false positives, or false negatives. In binary classification tasks involving disease pairs, the disease with the smaller sample size was defined as the positive class. The training/test splits maintained the original ratio of healthy to diseased samples.

Performance metrics included classification accuracy, sensitivity, specificity, and the area under the receiver operating characteristic curve for binary tasks. For multiclass (4-class) classification, overall accuracy was reported and visualized using a confusion matrix.

#### Visualization of Disease Interrelationships

To visualize the relationships among disease groups, we used the VOC combination that yielded the highest accuracy in 4-class classification. We first applied UMAP ^47^, a nonlinear dimensionality reduction method that preserves the topological structure of the original high-dimensional data in two dimensions. This enabled intuitive visualization of sample distribution across disease states.

Next, we applied PAGA ^16^, originally developed for single-cell transcriptomic analysis, to model state transitions between health conditions. In the PAGA graph, nodes represent clusters of samples, and edges represent similarity-based transitions between them.

To infer dynamic disease trajectories, we used CellRank ^17^, which estimates transition probabilities based on a pseudotime kernel. Clustering was performed using the Leiden algorithm, and a starting point (“root”) was defined in the cluster containing healthy participants. Transition paths were overlaid on the UMAP projection, illustrating plausible trajectories from health to various disease states.

#### Statistical analysis

To examine potential confounding by age and smoking status ^18^, correlation ratio analyses were conducted with VOC concentration as the dependent variable and age or smoking status (smoker vs. non-smoker) as independent variables. Additionally, we used the Kruskal–Wallis test to assess significant differences in diagnostic accuracy across VOC combinations identified via the wrapper method. A p-value < 0.01 was considered statistically significant. All analyses were performed using Python 3.8.

## Abbreviations

Ctrl: Healthy control participants
GCCMS: Gas chromatography–mass spectrometry
HBV: hepatitis B virus
HCC: Hepatocellular carcinoma
HCV: hepatitis C virus
MASH: Metabolic dysfunction-associated steatohepatitis
MASLD: Metabolic dysfunction-associated steatotic liver disease
NAFLD: Nonalcoholic fatty liver disease
NASH: Nonalcoholic steatohepatitis
PAGA: Partition-Based Graph Abstraction
PD: Periodontitis
PVDF: polyvinylidene fluoride
P450: Cytochrome P450
SVM: Support Vector Machine
UMAP: Uniform Manifold Approximation and Projection
VOCs: Volatile organic compounds

## Acknowledgements

The study team would like to sincerely thank all the participants. We thank Mr. H. Tokutake from Aichi Prefectural University, Professor J. Kato from Nara Institute of Science and Technology, and Dr. T. Itoh from the National Institute of Advanced Industrial Science and Technology (AIST) for their helpful discussions and technical comments.

## AUTHOR CONTRIBUTIONS

Conceptualization, K.T.S., Y.S., and M.M.; Formal Analysis, G.F., A.T., Y.S., K.I., and K.T.S.; Funding Acquisition, K.T.S., W.S., K.S., Y.S., and M.M.; Investigation, K.O., S.S., A.N., T. Takemoto, and H.H.; Resources, K.O., W.S., T. Takemoto, H.A., and H.H.; Software Programming, K.T.S. and Y.K.; Supervision, Y.S., K.I., and M.M.; Writing – Original Draft Preparation, K.T.S., Y.S., and M.M.; Writing – Review & Editing, Y.S., M.M., K.T.S., K.I., W.S., A.N., S.S., H.A., H.H., K.S., and T. Takata.

## DATA AVAILABILITY

The datasets generated and/or analyzed during the current study are not publicly available in accordance with the ethical guidelines set by the Japanese government for medical and biological research involving human participants.

