## Supplemental Information for "Trajectory-Informed Breathomics for Dynamic Mapping of Health and Disease: Toward a *Health Navigation Framework*"

### Supplemental figures and tables

**Table S1. Detection rate (%) of each VOC in healthy subjects and by disease.** Percentage of subjects whose samples were evaluated by GC/MS in this study.

| VOC | Control | NASH | HCC | PD |
|---|---|---|---|---|
| Acetic acid | 100 | 100 | 98 | 97 |
| Acetoin | 72 | 81 | 93 | 83 |
| Acetone | 100 | 100 | 100 | 100 |
| Butyric acid | 19 | 70 | 46 | 38 |
| p-cymene | 82 | 73 | 95 | 83 |
| Dimethyl sulfide | 98 | 97 | 99 | 93 |
| Hydrogen | 95 | 99 | 92 | 97 |
| Limonene | 100 | 100 | 100 | 100 |
| Methanethiol | 87 | 93 | 93 | 69 |
| Nonanal | 100 | 100 | 100 | 100 |
| Propionic acid | 8 | 85 | 86 | 90 |
| Styrene | 100 | 99 | 99 | 100 |
| Toluene | 100 | 100 | 100 | 100 |
| m-xylene | 100 | 100 | 100 | 100 |
| o-xylene | 100 | 100 | 100 | 100 |
| p-xylene | 100 | 100 | 100 | 100 |

**Table S2. Description of VOCs classified in Table 1 and their related disease(s).** IPF indicates idiopathic pulmonary fibrosis, and GERD indicates gastroesophageal reflux disease.

| VOC | Description | Biogenesis | Related disease(s) |
|---|---|---|---|
| p-cymene | Metabolized by the P450 with extrinsic factors. | × | cirrhotic CLD[1] |
| Limonene | Metabolized by the P450 enzymes CYP2C9 and CYP2C19 with extrinsic factors. | × | cirrhotic CLD[2], NASH[3], HCC[4] |
| Styrene | Metabolized by the P450 with extrinsic factors. | × | HCC[5], cirrhotic without HCC[6] |
| Toluene | Metabolized by the P450 with extrinsic factors. | × | Lung cancer[7] |
| (m, o, p)-xylene | Metabolized by the P450 with extrinsic factors. | × | Lung cancer[8] |
| Acetoin | Produced by *Lactococcus lactis*. | ✓ | IPF[9], Lung cancer[10], |
| Acetic acid | A type of short-chain fatty acid, produced by gut bacteria and alcohol metabolism in the liver. | ✓ | Decompensated cirrhosis[11], NAFLD[12], GERD[13], |
| Butyric acid | A type of short-chain fatty acid produced by gut bacteria. | ✓ | NAFLD[12] |
| Hydrogen | Produced by gut bacteria, $H_2$ in exhaled gas correlates with the amount of $H_2$ produced in the gut[14]. | ✓ | Noncirrhotic HCC[15] |
| Propionic acid | A type of short-chain fatty acid produced by gut bacteria. | ✓ | Decompensated cirrhosis[11], NAFLD[12], cirrhotic CLD[16] |
| Acetone | Produced by lipid metabolism. | ✓ | Diabetes[17], cirrhotic CLD[5,18] |
| Dimethyl sulfide | One of the causative agents of halitosis. | ✓ | PD[19], cirrhotic CLD[5,16], Cirrhosis[20] |
| Methanethiol | One of the causative agents of halitosis. | ✓ | PD[19], Cirrhosis[20] |
| Nonanal | Formed mainly by oxidative degradation of sebum (oleic acid). | ✓ | Lung cancer[21] |

**Table S3. Function types and parameter ranges for SVM optimization.**

| Parameter | Search range |
|---|---|
| **Kernel Functions** | Linear functions, cubic functions, Gaussian functions |
| $\sigma$ | [0.1:0.1:2] (values in increments of 0.1 between 0.1 and 2) |
| $C$ | [1000:1000:10000] (values between 1,000 and 10,000 in increments of 1,000) |

**Table S4. Cross-correlation between 16 VOC concentrations and age.** Factors with a cross-correlation coefficient with age greater than 0.35 or less than −0.35 are marked with a check mark (significance level of p=0.05).

| VOCs | Control | NASH | HCC | PD |
|---|---|---|---|---|
| Acetone | x | x | x | x |
| Dimethyl sulfide | x | x | x | x |
| Methanethiol | x | x | x | x |
| Nonanal | x | x | x | x |
| Acetic acid | x | x | x | x |
| Acetoin | x | x | x | x |
| Butyric acid | x | x | x | x |
| Hydrogen | x | x | x | x |
| Propionic acid | x | x | x | x |
| p-cymene | x | x | x | x |
| Limonene | x | x | x | x |
| Styrene | x | x | x | x |
| Toluene | x | x | x | x |
| m-xylene | x | x | x | x |
| o-xylene | ✓ | x | x | x |
| p-xylene | ✓ | x | x | x |

**Table S5. Highest accuracy and VOC combinations for healthy and disease binary classification.**

| Disease | Accuracy | Sensitivity | Specificity | AUC | VOCs |
|---|---|---|---|---|---|
| NASH | 0.994 | 1.00 | 0.990 | 0.998 | Limonene, Toluene, (m, p)-xylene, Acetic acid, Butyric acid, Hydrogen, Propionic acid, Dimethyl sulfide, Nonanal |
| HCC | 0.974 | 0.979 | 0.970 | 0.988 | p-cymene, Limonene, Styrene, Toluene, (o, p)-xylene, Acetoin, Propionic acid |
| PD | 0.938 | 0.793 | 0.980 | 0.860 | Styrene, Toluene, (o, p)-xylene, Acetoin, Butyric acid, Hydrogen, Dimethyl sulfide, Methanethiol |

**Table S6. Highest accuracy of binary classification achieved between diseases and the combination of VOCs.** D-1 was designated the negative class, and D-2 was the positive class.

| D-1 | D-2 | Accuracy | Sensitivity | Specificity | AUC | VOCs |
|---|---|---|---|---|---|---|
| HCC | NASH | 0.920 | 0.947 | 0.881 | 0.907 | Acetic acid, Acetoin, Hydrogen, Methanethiol, Toluene, (m, p)-xylene |
| HCC | PD | 0.976 | 0.989 | 0.931 | 0.979 | Acetic acid, Acetoin, Dimethyl sulfide, Limonene, (m, o, p)-xylene |
| NASH | PD | 0.969 | 0.985 | 0.931 | 0.933 | Methanethiol, Styrene, Toluene, (o, p)-xylene |

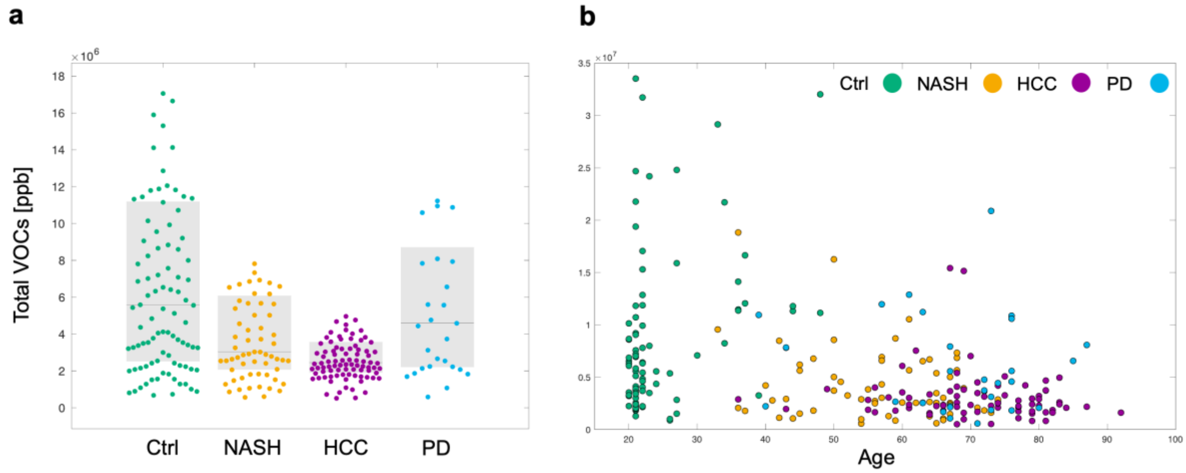


**Figure S1. Distribution of sums of VOCs in exhaled air per subject.** (a) Sums of VOCs for healthy (Ctrl) and diseased (NASH, HCC, PD) subjects, with 16 VOC concentrations measured for each subject (Total VOCs). Boxplots indicate median and interquartile range. (b) Total VOC concentrations by age, with subject status represented by the same color as in **a**.

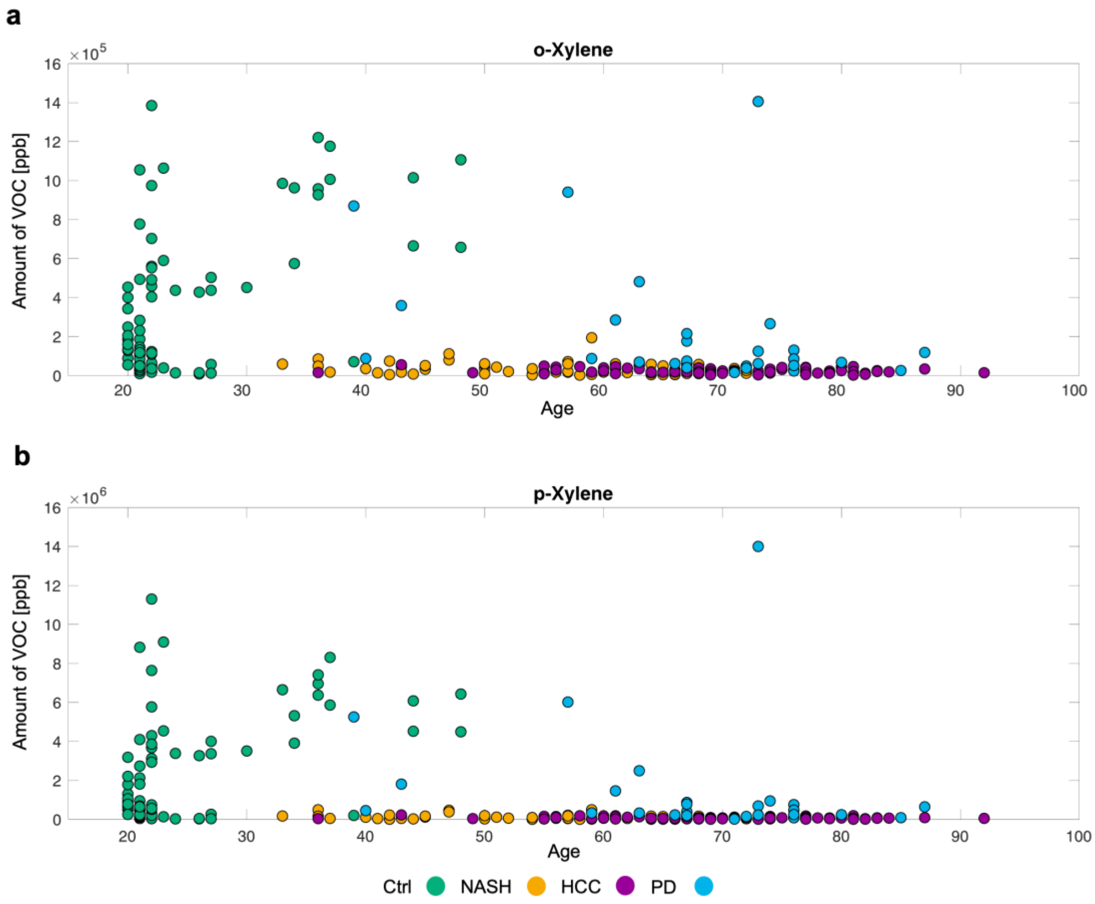


**Figure S2. Comparison of age and VOC levels by disease.** Comparison of (a) o-xylene and (b) p-xylene concentrations in healthy, NASH, HCC, and PD subjects.

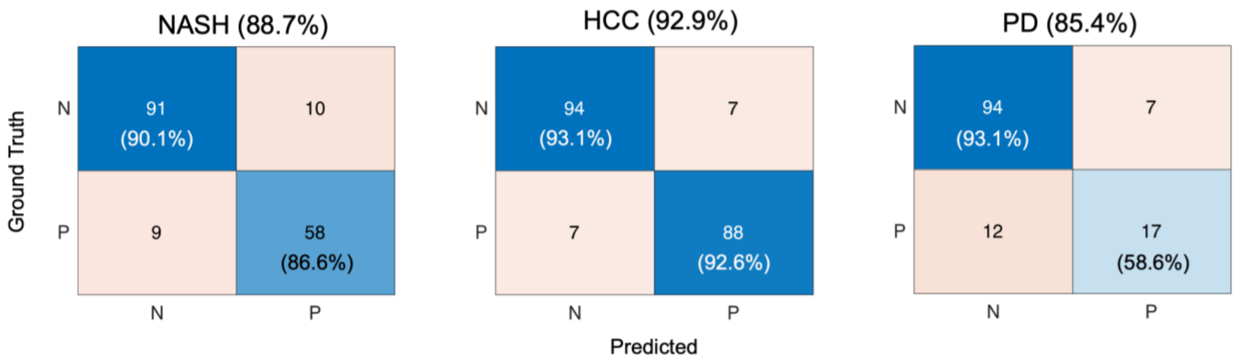


**Figure S3. Accuracy of binary classification learning by VOC concentration without preprocessing.** We conducted cross-validation for binary classification learning using the detected values as they were, without any concentration ratio transformation within the selected VOCs in the wrapper method.

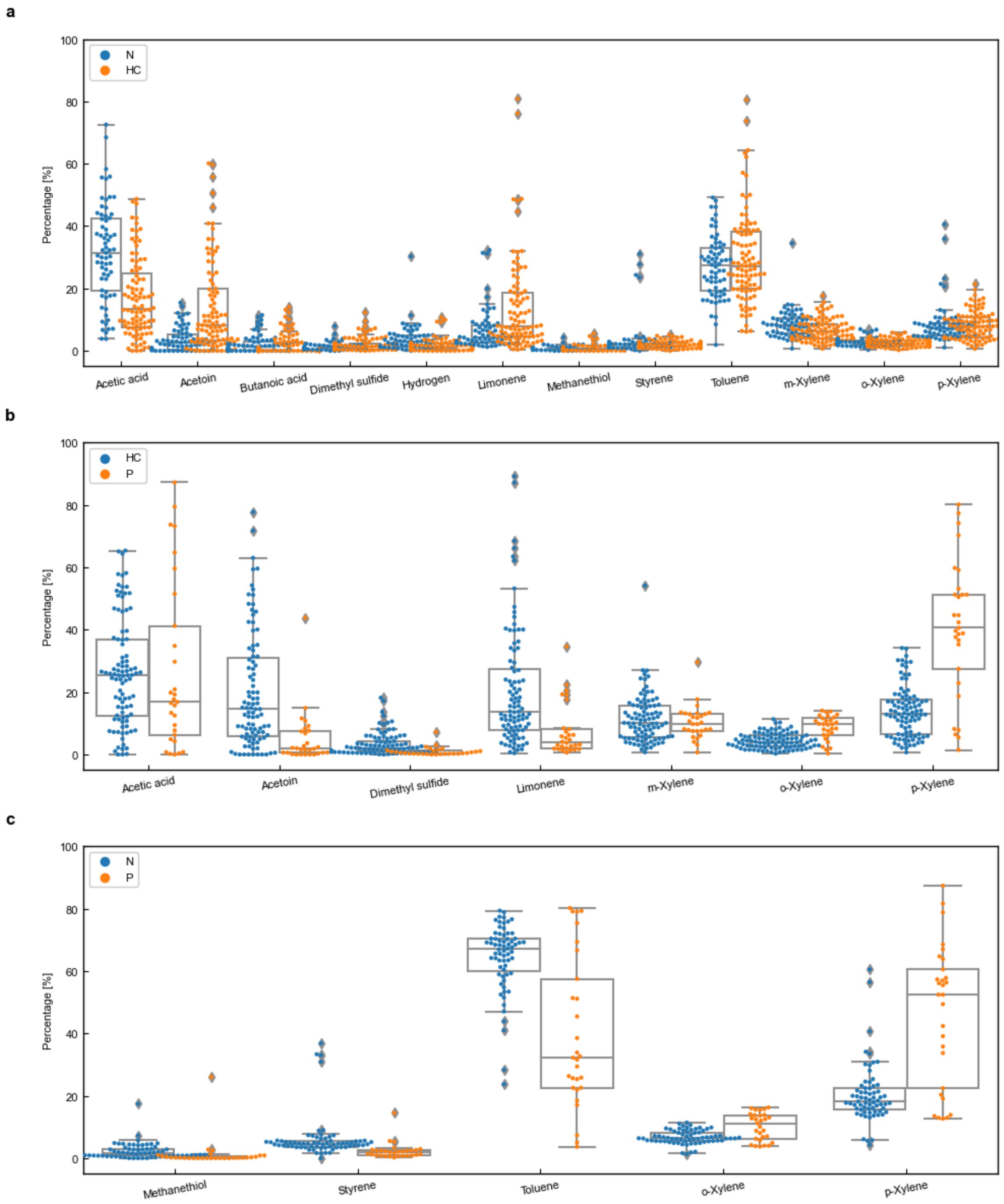


**Figure S4. Optimal VOC combinations for healthy vs. disease classification.**
Box plots show the distribution of VOC concentration ratios for the optimal combinations that achieved highest diagnostic accuracy in distinguishing (a) healthy controls from NASH patients, (b) healthy controls from HCC patients, and (c) healthy controls from PD patients. Blue dots represent healthy control participants (Ctrl), and orange dots represent disease group participants. The VOC combinations shown correspond to those listed in **Table S5** with the highest classification accuracy for each disease comparison. Box plots indicate median and interquartile range, with whiskers extending to 1.5 times the interquartile range. Individual data points are overlaid to show the distribution of values within each group. Abbreviations: Ctrl, healthy control; HCC, hepatocellular carcinoma; NASH, nonalcoholic steatohepatitis; PD, periodontitis; VOCs, volatile organic compounds.

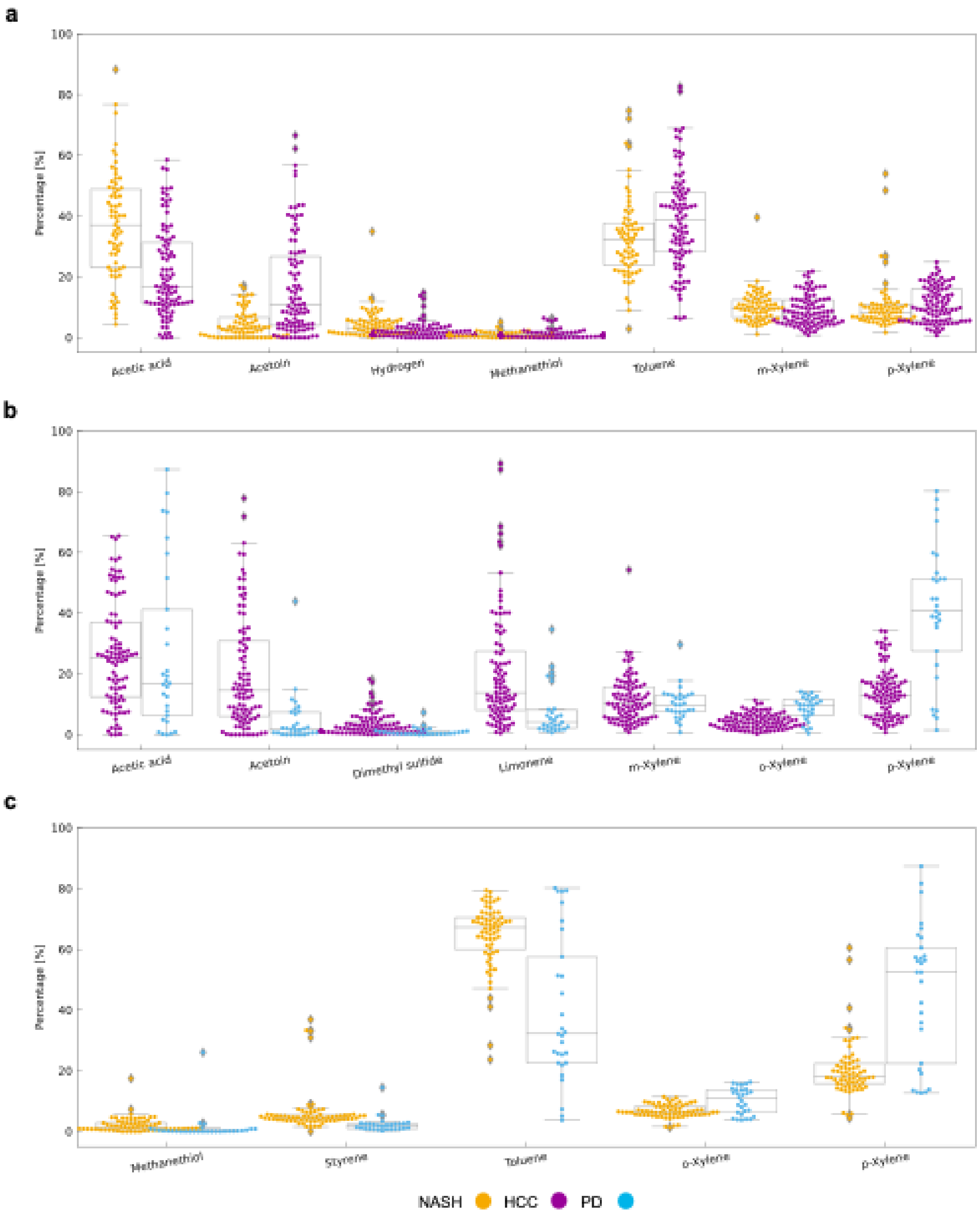


**Figure S5. Distribution of optimal VOC combinations for inter-disease classification.**
Box plots show the distribution of VOC concentration ratios for the optimal combinations that achieved highest diagnostic accuracy in distinguishing between different disease groups: (a) NASH versus HCC patients, (b) HCC versus PD patients, and (c) NASH versus PD patients. Each panel displays the VOC combinations that yielded the highest classification accuracy for the respective disease pair comparisons. Orange dots represent NASH patients, purple dots represent HCC patients, and blue dots represent PD patients. The VOC combinations shown correspond to those listed in **Table S6** with the highest classification accuracy for each inter-disease comparison. Box plots indicate median and interquartile range, with whiskers extending to 1.5 times the interquartile range. Individual data points are overlaid to show the distribution of values within each group. These results demonstrate disease-specific VOC signatures that enable differential diagnosis between different pathological

conditions. Abbreviations: HCC, hepatocellular carcinoma; NASH, nonalcoholic steatohepatitis; PD, periodontitis; VOCs, volatile organic compounds.
